# Prevalence and factors associated with undernutrition among under-five children with sickle cell disease at Mulago National Referral Hospital, Uganda: A facility-based cross-sectional study

**DOI:** 10.1101/2024.12.18.24319255

**Authors:** Annet Namubamba, Henry Wamani, Agnes Namaganda, Umusalima Namagala, Priscilla Cheputyo, Dan Muramuzi, Saul Kamukama

**Affiliations:** Department of Community Health and Behavioral Sciences, School of Public Health, College of Health Science, Makerere University; Department of Pharmacy, School of Health Sciences, College of Health Science, Makerere University; Department of Physiology, School of Biomedical Sciences, College of Health Science, Makerere University; Department of Paediatric and Child Health, School of Medicine, College of Health Science, Makerere University; Department of Epidemiology and Biostatistics, School of Public Health, Makerere University, Kampala, Uganda

**Keywords:** Sickle cell disease, Undernutrition, under-five, Children

## Abstract

**Background:** Sickle cell disease (SCD) significantly contributes to under-five morbidity and mortality in sub-Saharan Africa and increases vulnerability to undernutrition. However, limited studies exist on the prevalence of undernutrition and its associated factors among children under five years with SCD in Uganda. We assessed the prevalence of undernutrition and its associated factors among children with SCD aged 6-59 months attending the sickle cell clinic at Mulago National Referral Hospital in Kampala, Uganda.

**Methods:** A facility-based cross-sectional study was conducted among 329 children aged 6-59 months. Data was collected in August 2022 using a structured questionnaire and analyzed using STATA version 14.0. Modified Poisson regression was used to identify the factors associated with undernutrition.

**Results:** The prevalence of undernutrition among children with SCD aged 6-59 months was 31.3% (n=103) 95% CI 26.5, 36.5; 27.4% (n=90) 95% CI 22.8, 32.5 were stunted, and 14.3% (n=47) 95% CI 10.8, 18.5 were underweight. Undernutrition among children with SCD was statistically significantly associated with children below 24 months but not breastfeeding (aPR=1.27; 95% CI: 1.09-1.48; P-0.002), not immunized against measles (aPR=1.31; 95% CI: 1.02-1.68; P-0.03), and child not taking hydroxyurea drug (aPR=1.5; 95% CI: 1.07-2.08; P-0.017).

**Conclusion:** Early weaning of children, not being immunized against measles, and the child not taking hydroxyurea drug were great contributors to undernutrition. There is a need for sensitization of the caretakers about these predictors, and integrating hydroxyurea treatment in the standard care package of management of SCD in Uganda.

## BACKGROUND

Sickle cell disease also known as sickle cell anaemia is one of the common inherited blood disorders in humans characterized by producing defective haemoglobin leading to sickle-shaped red blood cells, and has been associated with a decreased dietary intake which results in poor nutrition status and impaired growth [1]. It is a lifetime disease and is characterized by recurrent episodes of pain. The recurrent pain results from sticky and stiff red blood cells that clog tiny blood vessels and cause infections, acute chest syndrome, priapism, and stroke in worse situations [2, 3]. The painful episodes cause acute illness which results in poor appetite. This limits nutrient intake; low nutrient intake causes weight loss, muscle wasting, growth faltering, and micronutrient deficiency which leads to impaired immunity response in people living with SCD rendering them vulnerable to increased infection and disease [4, 5]. This whole cycle leads to undernutrition like wasting, stunting, and underweight among people living with SCD. In SCD there is a continuous breakdown of sickle red blood cells called haemolyis, which results in anaemia [5, 6]. Anaemia is one of the micronutrient deficiencies that contributes to the mortality rate among people living with SCD. People living with SCD have a greater red blood cell turnover, increased energy requirements, and increased protein turnover [7, 8].

SCD in association with undernutrition significantly contributes to increased morbidity and mortality of children below 5 years of age [9]. Nutrition status has been implicated in increasing the severity of sickle cell disease [10]. Undernutrition (underweight and/or stunting) among these children leads to delayed growth and development; and retarded mental development. This causes poor performance in school and reduced productivity in society hence proving to be a public health burden [5, 7, 8, 11].

The National Sickle Cell survey 2015 showed that 13% of Ugandans had sickle cell trait of which 0.7% had sickle cell disease. In Uganda, 20,000 babies are estimated to be born with SCD of which 50% to 90% die before their 5^th^ birthday due to infections resulting from underlying poor nutrition status [3, 12, 13]. The burden of SCD is national and found in all regions of Uganda. The burden of SCD in Uganda is highest in the East Central region (1.5%), Mid Northern region (1.3%), and Mid-Eastern region (1.2%); and lowest in the Southern West region (0.2%). The burden of SCD in Kampala alone a district found in the central region is 0.7% [12]There are about 10 sickle cell clinics in the central region of Uganda, the first of which was the Mulago Sickle Cell Clinic at Mulago National Referral Hospital. It serves the biggest number of SCD patients in the central region. However, being in a National referral Hospital also serves SCD patients referred from the nearby regions.

The Ministry of Health has made significant strides in addressing the sickle disease burden by coming up with National Guidelines for Management and Prevention of Sickle Cell Disease (which was launched in 2020 during the commemoration of World Sickle Cell Day), introducing free newborn screening programs in selected districts with the highest disease burden [14]. There have been efforts for community awareness and prevention of SCD by NGOs like Uganda Sickle Cell Rescue Foundation; and by cultural leaders like the previous Kabaka of Buganda birthday runs aimed at sickle cell awareness and screening in the region [13]. All interventions are aimed at awareness, screening, management of outcomes, increasing care, and reducing morbidity and mortality. However, nutrition care for SCD is not amply addressed in all the interventions available. Yet, undernutrition is one of the underlying factors that significantly contribute to the increased morbidity and mortality among children with SCD below five years of age in Uganda. Therefore, this justified this study by providing the missing link in data on undernutrition among children below 5 years of age and its associated factors. This information can be used to improve nutrition screening and advocate for nutrition interventions to be included in the routine management of SCD.

## METHODS

### Study Design and Study Setting

A facility-based cross-sectional study design was used. The study was conducted at the sickle cell clinic at Mulago National Referral Hospital (MNRH), which is located on Mulago Hill at coordinates 0°20’27”N, 32°34^’^33.4”E. This clinic serves a population of about 800 individuals with SCD, predominantly living in the central region of Uganda. The clinic operates five days a week (Mon-Fri) serving 50-80 clients a day.

### Study Population and Eligibility Criteria

The study population was children of 6-59 months diagnosed with sickle cell disease by *HbSS* (Haemoglobin S) positive and receiving routine care at the outpatient sickle cell clinic (SCC) at MNRH. The diagnosis would be cross-checked from the patient’s file. Children who were in crises and needed hospitalization, children whose caretakers were less than 18 years of age, children with chronic co-morbidities including tuberculosis and kidney failure; and children with disabilities were excluded.

### Study variables

The dependent variable was undernutrition (underweight and/or stunting) and this was measured as having <-2SD Z-scores for the weight-for-age and/or height-for-age anthropometric indices from the reference population [6].

The independent variables were categorized into child characteristics and caretaker/household characteristics and were measured using a standardised questionnaire adapted and modified from Kasajja et al, 2021 and Nsubuga et al, 2020 [15, 16]. The child’s characteristics included: age (in months), sex, birth order, disease history in the past two weeks, liver or kidney disease, prophylaxis taken (penicillin or hydroxyurea), and food intake. A validated dietary diversity score based on the 12 food groups consumed in the past 24 hours before the study was used to assess food intake. A score of one or zero was awarded for each of the food groups consumed or not consumed respectively [17]. The caretaker/household characteristics included; age (years), marital status, religious affiliation, highest level of education, occupation, number of children in the home, sex of head of family, and household size.

### Sample size and sampling procedure

This study adopted the Kish Leslie (1965) formula for sample size calculation for cross-sectional studies[19]. The total sample size was 329, and the key parameter used was Where q=(1-p), a Z-value corresponding to a 95% level of confidence at 1.96, using the prevalence of stunting at 25.8% [20], and a 5% level of precision was used to calculate the sample size for the study. Consecutive sampling was used to sample the participants.

### Data Collection Procedure

Data was collected from 8th August 2022 and ended on 25th August, 2022 using face-to-face interviews with a structured questionnaire by trained research assistants. The questionnaire was translated to Luganda which was the commonly spoken language in the study area. Questionnaires were filled to completion and submitted to the principal investigator for cross-checking. The weight was measured using standard techniques as stipulated by WHO standards. Seca 877 flat weighing scale was calibrated to zero before taking every measurement. The children were weighed in standing position with minimum clothing and without shoes; for children who cannot stand, weight was taken by using the 2 in 1 option with the caregiver carrying the infant with minimal clothes. Weight was recorded in kilograms to the nearest 0.1kg. This was done twice so that an average weight was calculated. The procedures were obtained from the reference manual for Integrating Nutrition Assessment, Counseling, and Support into Health Service Delivery [11]. Measurement of height for children less than 2 years was taken in the lying position using seca 417 portable infantometer with the aid of the caregiver and measurement of height for children 2 years and above was taken in the standing position using seca 213 portable stadiometer. Length/height was taken in centimeters to the nearest 0.1cm. This was done twice so that an average height was calculated [11].

### Data analysis

Collected data was entered into an Excel spreadsheet, cleaned, and then exported to STATA software version 14.0 (Texas USA) for analysis. Social demographic data was summarized using descriptive statistics; frequencies and percentages for categorical variables and mean (standard deviation), median (IQR) for continuous variables. The children’s age was converted to months and anthropometric measures of weight and height was used to measure underweight and stunting respectively. Weight-for-age (WAZ) and Height-for-age (HAZ) Z-scores were computed using the WHO Child growth reference standards [21]. Normal nutrition status was defined as Z-scores ≥-2SD, underweight and stunting were defined as Z-scores <-2SD and dichotomously categorized as yes/no. The prevalence of underweight and stunting was expressed in percentages.

At bivariable analysis, the factors associated with undernutrition were determined using a modified Poisson regression. The resulting crude prevalence ratios (cPR) and their corresponding 95% confidence interval (CI) were interpreted using a p-value of <0.2. Independent variables with p-value <0.2 were selected for multivariable analysis. Additionally, independent variables with p-values > 0.2 that were essential to the study based on literature and clinical significance (dietary intake and sanitation practices) were also considered for multivariable analysis. In multivariable analysis, the stepwise regression method was used in model building. Interaction between variables was assessed using the Wald test and no interaction term was statistically significant. Collinearity between variables was assessed and no correlation was found between any of the selected variables. The results of the multivariable analysis were presented as Adjusted Prevalence ratios (aPR) and their corresponding p-values at 95% CI.

### Ethical approval

Ethical clearance was obtained from Makerere University School of Public Health Higher Degrees, Research and Ethics Committee (MakSPH-REC 62). An introductory letter from the School of Public Health was used to seek administrative clearance from the Executive Director of MNRH. Written informed consent was obtained from the parents/caretaker.

## RESULTS

### Respondent Characteristics

A total of 329 parents/caretakers participated in the study. The mean age of parents/caretakers was 31 years (± 7.33). The majority of parents/caretakers (73.3%) were married and (47.4%) attained a secondary level of education (Table 1).

**Table 1:** Characteristics of respondents of 329 children aged 6-59 months with SCD at MNRH enrolled in the study.

| Variable | Categories | Frequency (N=329) | Percentages (%) |
| --- | --- | --- | --- |
| Age (Years) | Median (IQR) | 31 (27, 36) |  |
| Marital status | Married | 241 | 73.3 |
|  | Not married | 88 | 26.7 |
| Relationship with child | Mother | 268 | 81.5 |
|  | Father | 28 | 8.5 |
|  | Others | 33 | 10.0 |
| **Education level** | Primary level or less | 58 | 17.6 |
|  | Secondary level | 156 | 47.4 |
|  | Tertiary level | 115 | 35.0 |
| **Occupation** | Farmer | 16 | 4.9 |
|  | Formal employment | 52 | 15.8 |
|  | Informal employment | 24 | 7.3 |
|  | Self-employed | 144 | 43.8 |
|  | Unemployed | 61 | 18.5 |
|  | Other | 32 | 9.7 |
| **Household members** | **Under 5 years of age** |  |  |
|  | <2 members | 193 | 58.7 |
|  | 2-4 members | 134 | 40.7 |
|  | >4 members | 2 | 0.6 |
|  | **Above 5 years of age** |  |  |
|  | <2members | 42 | 12.7 |
|  | 2-4 members | 216 | 65.7 |
|  | >4 | 71 | 21.6 |
| **Residence(region)** | Central | 309 | 94.0 |
|  | Eastern | 19 | 6.0 |
|  | Western | 1 | 0.0 |
| **Sex of household head** | Male | 238 | 72.3 |
|  | Female | 91 | 27.7 |
| **Individual that feeds the child** | Child feeds self | 144 | 43.7 |
|  | Parent | 159 | 48.3 |
|  | Others | 26 | 8.0 |
| **Have a toilet/ latrine at home** | Yes | 327 | 99.4 |
|  | No | 2 | 0.6 |
| **Have a hand washing facility at home** | Yes | 241 | 73.3 |
|  | No | 88 | 26.7 |
| **Have a garbage pit at home** | Yes | 269 | 81.8 |
|  | No | 60 | 18.2 |

Almost all respondents (n=327) had a toilet/latrine a home, however, 2 respondents did not have a toilet/latrine at home.

### Study Population Characteristics

The mean age of children aged 6-59 months with SCD at MNRH enrolled in the study was 31 months (SD 15.7) (Table 3). The study enrolled 171 (52.0%) male and 158 (48.0%) female children aged 6-59 months with SCD at MNRH (Table 2).

**Table 2:** Characteristics of 329 study children aged 6-59 months with SCD at MNRH.

| **Variable** | **Categories** | **Frequency (N=329)** | **Percentages (%)** |
| --- | --- | --- | --- |
| **Age group (months)** | 6-11 | 25 | 7.6 |
|  | 12-23 | 69 | 21 |
|  | 24-35 | 81 | 24.6 |
|  | 36-59 | 154 | 46.8 |
| **Sex of the child** | Male | 171 | 52.0 |
|  | Female | 158 | 48.0 |
| **Nutrition status of children** | Normal | 226 | 68.7 |
|  | Undernourished (WAZ <- 2 Z-Scores and/or HAZ <-2Z-scores) | 103 | 31.3 |
| **Had exclusive breastfeeding for at least 6 Months** | Yes | 214 | 65.1 |
|  | No | 115 | 34.9 |
| **Initiation of solid feeds** | 0-5Months | 102 | 31.0 |
|  | 6 Months and above | 227 | 69.0 |
| **Still breastfeeding** | Yes | 55 | 58.5 |
|  | No | 39 | 41.5 |
| **Number of meals taken by a child in a day** | Inadequate (<3) | 27 | 8.2 |
| | Adequate ($\geq 3$) | 302 | 91.8 |
| **No. of food groups consumed** | Inadequate (<4) | 35 | 10.6 |
|  | Adequate (4-6) | 163 | 49.5 |
|  | High (>6) | 131 | 39.9 |
| **Child dewormed in past 6 months (for age $\geq 12$ months)** | Yes | 243 | 79.9 |
|  | No | 61 | 20.1 |
| **Child received Vitamin A in the past 6 months** | Yes | 133 | 40.4 |
|  | No | 196 | 59.6 |
| **Child immunized against measles (for age $\geq 9$ months)** | Yes | 306 | 96.5 |
|  | No | 11 | 3.5 |
| **Had an illness in the past 2 weeks** | Yes | 113 | 34.4 |
|  | No | 216 | 65.6 |
| **Have a chronic disease other than SCD** | Yes | 8 | 2.4 |
|  | No | 321 | 97.6 |
| **Had malaria prophylaxis** | Yes | 226 | 68.7 |
|  | No | 103 | 31.3 |
| **Had penicillin prophylaxis** | Yes | 285 | 86.6 |
|  | No | 44 | 13.4 |
| **Child taking hydroxyurea drug** | Yes | 163 | 49.5 |
|  | No | 166 | 50.5 |

**Table 3:** Multivariate analysis of factors associated with under-nutrition among 329 children with SCD in MNRH.

| Variable | Categories | Prevalence of undernutrition (n=103) (%) | cPR | 95% CI | P- value | aPR | 95%CI | P- Value |
| --- | --- | --- | --- | --- | --- | --- | --- | --- |
| **Age (Months)** | Median (IQR) |  | 0.99 | 0.98, 1.00 | 0.045 |  |  |  |
| **Age groups (months)** | 6-11 | 2(1.9) | 1 |  |  |  |  |  |
|  | 12-23 | 31(30.1) | 1.34 | 1.18,1.52 | <0.001 |  |  |  |
|  | 24-35 | 33(32.1) | 1.30 | 1.15,1.48 | <0.001 |  |  |  |
|  | 36-59 | 37(35.9) | 1.15 | 1.03,1.29 | 0.02 |  |  |  |
| **Sex of the child** | Female | 41(25.9) | 1 |  |  |  |  |  |
|  | Male | 62(36.3) | 1.47 | 1.31,1.66 | 0.00 |  |  |  |
| **Had exclusive breastfeeding for at least 6 Months** | Yes | 61 (28.5) | 1 |  |  |  |  |  |
|  | No | 42 (36.5) | 1.28 | 0.93,1.77 | 0.131 |  |  |  |
| **Initiation of solid feeds** | 0-5Months | 39 (38.2) | 1 |  |  |  |  |  |
|  | 6 Months and above | 64 (28.2) | 0.74 | 0.53, 1.02 | 0.064 |  |  |  |
| **Child still breastfeeding (for ≤23months)** | Yes | 11(20.0) | 1 |  |  | 1 |  |  |
|  | No | 22(56.4) | 1.30 | 1.14,1.44 | <0.001 | 1.27 | 1.09,1.48 | **0.002** |
| **Number of food groups consumed** | Adequate (4-5) | 59(57.3) | 1 |  |  |  |  |  |
|  | Inadequate (<4) | 8(7.8) | 0.9 | 0.80,1.02 | 0.12 |  |  |  |
|  | High (≥6) | 36(35.0) | 0.93 | 0.86,1.01 | 0.12 |  |  |  |
| **Marital status of parent/caretaker** | Married | 66(27.4) | 1 |  |  | 1 |  |  |
|  | Not married | 37(42.1) | 1.54 | 1.11,2.12 | 0.009 | 0.95 | 0.79,1.15 | 0.614 |
| **Relationship with child** | Mother | 79(29.5) | 1 |  |  |  |  |  |
|  | Father | 12(42.9) | 1.45 | 0.91,2.32 | 0.116 |  |  |  |
|  | Others | 12(36.4) | 1.23 | 0.76,2.01 | 0.400 |  |  |  |
| **Education level of parent/caretaker** | Primary and below | 30(26.1) | 1 |  |  |  |  |  |
|  | Secondary | 24(41.4) | 1.59 | 1.03,2.45 | 0.038 |  |  |  |
|  | Tertiary | 49(31.4) | 1.20 | 0.82,1.77 | 0.346 |  |  |  |
| **Had an illness in the past 2 weeks** | Yes | 30 (26.6) | 1.27 | 0.89, 1.82 | 0.188 |  |  |  |
|  | No | 73 (33.8) | 1 |  |  |  |  |  |
| **Immunized against measles (for age ≥ 9 months)** | Yes | 95(31.5) | 1 |  |  | 1 |  |  |
|  | No | 6(54.6) | 1.18 | 0.97,1.43 | 0.10 | 1.31 | 1.02,1.68 | **0.03** |
| **Had hydroxyurea** | Yes | 40 (24.5) | 1 |  |  | 1 |  |  |
|  | No | 63 (38.0) | 1.55 | 1.11, 2.16 | 0.010 | 1.5 | 1.07,2.08 | **0.017** |

The majority of the children (n=214, 65.1%) were exclusively breastfed (EBF) for the first 6 months of their life. However, 3 children were never breastfed at all among all the 329 children enrolled in the study. Up to 49.5% (n=163) of children with SCD enrolled in the study were taking the hydroxyurea. The majority of children (n=306, 96.5%) aged ≥9 months were immunized against measles.

### Prevalence of undernutrition among children aged 6-59 months with SCD at MNRH enrolled in this study

The overall prevalence of undernutrition among children aged 6-59 months with SCD at MNRH enrolled in this study was 31.3% (n=103). The prevalence of undernutrition was higher among boys (n=62, 36.3%) than in girls (n=41, 25.9%). In this study, the prevalence of undernutrition increased with increasing age and was highest in the age group of 36-59 months (n=37, 35.9%). Figure 2 below shows the prevalence of undernutrition in the study population.

**Figure 1:**
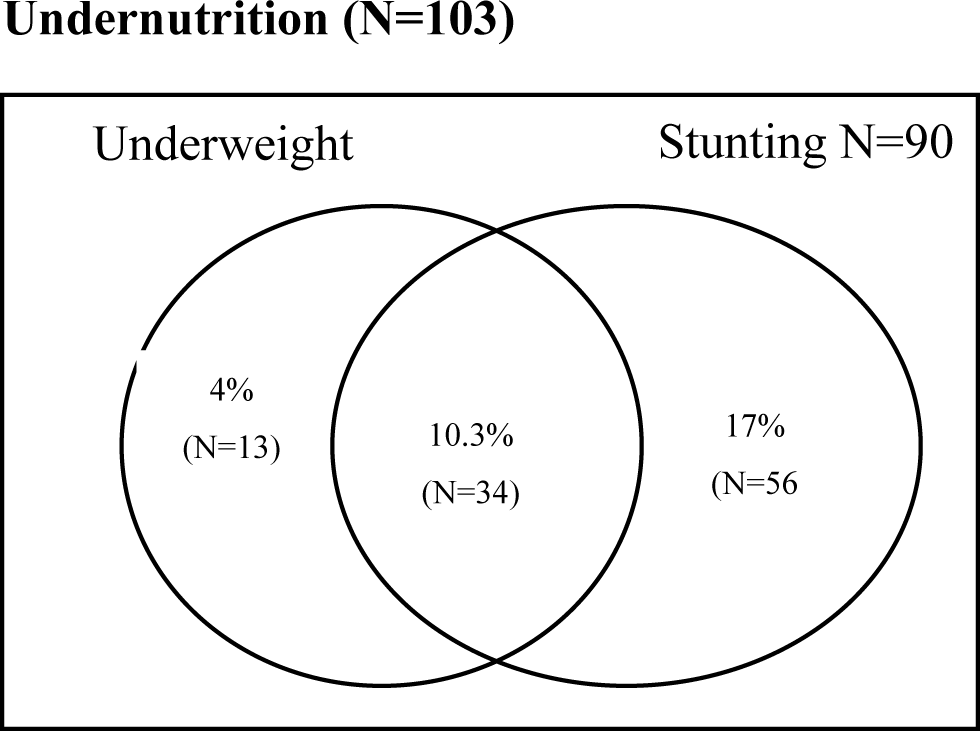
A Venn diagram showing the prevalence of undernutrition of children aged 6-59 months with SCD at MNRH. The prevalence of underweight and stunting was 14.3% (n=47) and 27.4% (n=90) respectively (fig.1)

In this study, 4% (n=13) of the children presented with only underweight, 10.3% (n=34) presented with both underweight and stunting; and 17% (n=56) presented with only stunting as shown in Figure 1 above.

### Multivariate analysis of factors associated with under-nutrition among 329 children with SCD in MNRH

The prevalence of undernutrition among children who were not taking hydroxyurea was 1.5 times (aPR 1.5; CI 1.07-2.08; P-0.017) compared to that of children taking hydroxyurea after controlling for other independent variables. The prevalence of undernutrition among children who were not immunized against measles was 1.31 times (aPR 1.31; 95% CI 1.02,1.68; P-0.003) compared to those immunized against measles. The prevalence of undernutrition among children who had discontinued BF before 24 months of age was 1.27 times (aPR 1.27;95% CI 1.09,1.48; P-0.002) compared to children who were still BF (Table 1).

## DISCUSSION

This study aimed to determine the prevalence of undernutrition and its associated factors among children with SCD aged 6-59 months at Mulago National Referral Hospital, Kampala, Uganda. It was a facility-based cross-sectional study that involved a total of 329 children with SCD attending the Outpatient Sickle Cell Clinic of MNRH. The prevalence of undernutrition in this study was 31.3% (n=103) and the factors found to be significantly associated with undernutrition were, discontinuation of breastfeeding before 24 months of age, a child not being immunized against measles, and not taking hydroxyurea drug.

### Prevalence of Undernutrition among children with SCD aged 6-59 months in MNRH

The overall prevalence of undernutrition among children enrolled in the study was 31.3%. In this study, the prevalence of stunting and underweight was higher than the national prevalence value of stunting (25%) and underweight (7.8%) as reported in the Nutrition Situation Report of the Uganda National Panel Survey 2019/20 [22]. This could be because children with SCD have slowed growth as compared to normal children who are included in the national panel survey [23]. In contrast, a study conducted in Nigeria reported a higher prevalence of stunting (55.4%) and underweight (38.9%) in children with SCD below 5 years of age compared to this study [5]. This big difference may be attributed to the study setting, that is to say, the current study was done in an urban setting while the Nigeria study was in a rural setting. This could be attributed to the good health-seeking behavior of people in urban areas, and having small families in urban areas hence less competition for food [24, 25]. Another descriptive cross-sectional study conducted in a rural setting in Rio de Janeiro, Brazil also reported a higher prevalence of stunting(35.1%) and underweight (16.2%) among children with SCD [26].

Undernutrition was more prevalent among boys (36.3%) than in girls (25.9%), a finding similar to that reported in other studies conducted in sub-Saharan African countries and India [16, 24, 27]. In Sub-Saharan Africa, it has been highlighted that male children are more likely to become undernourished than females [25]. This could be because boys spend most of their time outdoor unlike girls who spend most time indoors and the fact that boys have a higher energy requirement than girls [20, 24]. It is also hypothesized that stunting in boys is associated with sex-specific hormones such as testosterone, luteinizing hormone, and follicle-stimulating hormone (FSH). FSH in boys disappears after 6 months which causes stunting whereas it stays high in girls until 3-4 years [28]. However, a cross-section study conducted in Accra and Kenya [20] reported no statistically significant difference in the prevalence of undernutrition between boys and girls [20, 29]. In this study, the prevalence of undernutrition increased with increasing age and was highest in the 36-59 months age group, similar results were reported in a cross-sectional household survey study done in South Sudan and a cross-sectional study done at Mulago among children with SCD aged 5-12years [23, 24]. This could be attributed to the fact that undernutrition (especially stunting) becomes more obvious after 24 months of age. As children grow, they have increased nutrient demand, they learn to feed themselves and therefore can easily be exposed to food-borne pathogens yet younger children may be protected by mothers’ immunity passed on through breast milk [24, 27].

### Factors associated with undernutrition among children with SCD aged 6-59 months

Children who were not taking hydroxyurea drug were 50% more likely to be undernourished than children taking hydroxyurea. A study done at Mulago Hospital reported similar results of improved nutrition status for children with SCD taking hydroxyurea drug [30]. The results could be attributed to the benefits of hydroxyurea. Hydroxyurea increases fetal hemoglobin levels which improves growth in children with SCD and reduces the occurrences of crises in children with SCD, which are associated with poor nutrition status[30, 31].

Children who were discontinued from breastfeeding before 24 months of age were 73% more likely to be undernourished than those still breastfeeding. Similar findings of statistical significance of an association between breastfeeding and undernutrition were reported in studies conducted in Ethiopia, India, and Myanmar[32–34]. The results are mostly attributed to the benefits of breast milk. Breast milk promotes growth and development; and boosts immunity which reduces the risk of morbidity[11]. Therefore, discontinuation of breastfeeding denies the child of breast milk benefits hence higher odds of the child becoming undernourished.

Children who were not immunized against measles had a higher likelihood of being undernourished (69%) than children immunized against measles. A multistage stratified random cluster sampling study conducted in Tokyo, Japan reported similar results of children not being immunized associated with undernutrition [35]. The results could be because not immunizing a child puts him/her at risk of morbidity which eventually causes undernutrition.

### Limitations of the study

The study did not consider seasonal variations especially the crises that warrant hospitalisation of children with SCD for specialized treatment. This study was conducted at an established SCD clinic within a referral hospital where patients and their caretakers have been empowered as routine clients. The findings of this study may not reflect the situation in the community or even in remote areas of Uganda.

### Generalizability of results

Findings from this study can only be generalized to children with SCD aged 6-59 months in a health facility setting. This is because the sampling frame is only representative of a health facility setting and not the all community.

### Conclusion

The prevalence of undernutrition in this study was 31.3% (n=103) and the factors found to be significantly associated with undernutrition were, discontinuation of breastfeeding before 24 months of age, a child not being immunized against measles, and not taking hydroxyurea drug. There is a need for sensitization of the public about the benefits of breastfeeding children till 24 months of age, immunization and hydroxyurea drug among SCD children; and integrating hydroxyurea drug in the standard care package of management of SCD in Uganda.

## Data Availability

information will only be available after acceptance,

## Acknowledgments

We highly appreciate the research participants and research assistants for participating in the study. We are greatly indebted to the staff of the School of Public Health, Makerere University for their invaluable support. We extend our gratitude to the staff of Sickle Cell Clinic Mulago National Referral Hospital for the assistance and hospitality offered during data collection.

## Availability of data and materials

The data and materials will be availed on request from the corresponding author.

## Funding

This study was not funded.

## Authors’ contribution

**Conceptualization:** Annet Namubamba, Umusalima Namagala, Cheputyo Priscilla, Agnes Namaganda

**Data curation:** Henry Wamani, Dan Muramuzi, Annet Namubamba, Saul Kamukama

**Format analysis:** Dan Muramuzi, Cheputyo Priscilla, Annet Namubamba

**Fund acquisition:** Annet Namubamba

**Investigation:** Annet Namubamba, Umusalima Namagala

**Methodology:** Dan Muramuzi, Annet Namubamba, Cheputyo Priscilla, Agnes Namaganda

**Project administration:** Annet Namubamba

**Resources:** Annet Namubamba

**Software:** Dan Muramuzi

**Supervision:** Henry Wamani, Saul Kamukama, Dan Muramuzi, Agnes Namaganda

**Validation:** Henry Wamani, Dan Muramuzi, Annet Namubamba

**Visualization:** Annet Namubamba, Dan Muramuzi

**Writing - original draft:** Annet Namubamba

**Writing- reviewing & editing:** Henry Wamani, Saul Kamukama, Agnes Namaganda, Dan Muramuzi, Priscilla Cheputyo, Umusalima Namagala

## Notes

### Competing Interest Statement

The authors have declared no competing interest.

### Funding Statement

The author(s) received no specific funding for this work.

### Author Declarations

Ethical clearance was obtained from Makerere University School of Public Health Higher Degrees, Research and Ethics Committee (MakSPH-REC 62).

